# Exploring Stakeholder Support for Student-Run Free Clinics to Address Gaps in Non-Communicable Disease Care in Ghana: An Exploratory Sequential Mixed-Methods Study

**DOI:** 10.1101/2025.08.06.25333122

**Authors:** Brian Amu Fleischer, Esi Berkoh, Bismark Amoh, Afriyie Badu, Derek Anamaale Tuoyire, Jeremy I Schwartz

**Author notes:** Correspondence: **E-mail:** **(BAF)**.

## Abstract

**Background**

Non-communicable diseases (NCDs), such as hypertension and diabetes, are leading contributors to morbidity and mortality in Ghana. Underserved communities face persistent barriers to NCD screening, early management, and self-care education, limiting progress in disease control and health equity. Student-run free clinics (SRFCs) have successfully addressed similar gaps in healthcare access in other global contexts, but their feasibility and relevance remain underexplored in sub-Saharan Africa. This study investigates local stakeholder perspectives and support for SRFCs as a contextually appropriate intervention to expand access to NCD care in Ghana.

**Methods**

We used an exploratory sequential mixed-methods design with interpretation-level integration. Qualitative data were collected through six focus group discussions (FGDs) with 48 health professional students, six key informant interviews (KIIs) with faculty, deans, and Ghana Health Service (GHS) officials, and two FGDs with 12 community members. Quantitative data from surveys of 316 students assessed interest in SRFC participation and its predictors. Thematic analysis, descriptive statistics, and logistic regression were used for qualitative and quantitative data, respectively. Findings were integrated narratively, weaving together quantitative results and qualitative insights for convergence and contextual depth.

**Results**

Stakeholders across all groups—including health professional students, university faculty and leadership, GHS officials, and community members—expressed strong support for student-led initiatives to address gaps in NCD care. Integrated findings revealed three key domains: (1) widespread familiarity among students with NCDs and barriers to care; (2) strong student acceptance of SRFCs as a relevant, feasible, and impactful intervention to improve NCD screening, early management, and self-care education—shaped by both motivating and discouraging factors such as social impact, academic obligations, resource constraints, and safety concerns; and (3) cross-sector endorsement of SRFCs by institutional and community stakeholders, who emphasized alignment with organizational missions, confidence in student capacity, and the initiative’s value for hands-on training and interprofessional collaboration. Quantitatively, 86.7% of surveyed students expressed willingness to participate in SRFCs, and greater familiarity with NCDs was a significant predictor of interest (OR = 1.41, p = 0.007). Stakeholders viewed SRFCs as timely, trusted, and well-positioned to complement existing services such as GHS Wellness Clinics.

**Conclusions**

SRFCs represent a promising model to enhance NCD care in Ghana. Broad stakeholder support, institutional alignment, and a foundation of student-led health initiatives suggest strong feasibility for implementation. Future efforts should focus on structured supervision, resource planning, and sustainable integration with national health systems to maximize impact and scalability.

## Introduction

Student-run free clinics (SFRCs) have emerged as crucial safety nets delivering essential primary medical care to underserved populations (1,2). Staffed by dedicated students of medicine, nursing, pharmacy, and other health professional programs (herein referred to as “health professional students”) under the supervision of faculty, these clinics help address healthcare disparities and improve public health. In South Africa, the Students’ Health and Welfare Centers Organization (SHAWCO) operates over 200 clinics with more than 1,250 student volunteers across eight provinces (2). In the United States, over 140,000 patients are seen annually at 208 SRFC sites, with at least 75% of accredited medical schools supporting them (3).

In Ghana, non-communicable diseases (NCDs) pose a significant public health challenge. Hypertension, affecting 34% of Ghanaians, is the leading cause of death and the third leading cause of hospital admissions, accounting for 15.3% of total deaths and 4.7% of hospitalizations (4,5). Despite this high disease burden, Ghana faces a severe shortage of healthcare providers, with fewer than one physician per 10,000 people (6). This imbalance between healthcare needs and available resources underscores the urgency for innovative and sustainable interventions. Student-led initiatives, particularly within SRFCs, offer a promising approach to addressing these gaps by extending healthcare access and providing early screening, management, and education for NCDs.

Integrating cost-effective NCD interventions in primary care has been shown to be effective in reversing disease progression, preventing complications, reducing hospitalizations, lowering healthcare costs (7). Task-shifting, which delegates specific medical responsibilities from doctors to trained non-physician healthcare workers (NPHWs) and community health officers (CHOs), has proven effective in areas with physician shortages (8). Ghana’s Community-based Health Planning and Services (CHPS) program, originally designed for maternal and child health, exemplifies this approach (8). Despite the program’s success in transforming maternal and child health over the past 20 years, it lacks the training, medications, and resources necessary to address NCDs. As such, it remains poorly equipped to respond to the NCD epidemic, thereby justifying innovative solutions that build on the current model and other cost-effective interventions (9).

Interprofessional SRFCs has proven to be instrumental in delivering comprehensive care to underserved communities (10). They create an environment of collaboration, continuous learning, and skill development (11). Ghana’s health education system offers a rich pool of expertise across medical, pharmacy, nursing, public health, and allied health disciplines (12). Medical students bring diagnostic and clinical expertise for NCD screening and management. Nursing students offer patient care and counseling to support self-management. Pharmacy students ensure proper medication management and adherence. Public health students contribute community health promotion and disease prevention strategies. Other allied health students, including those in social work, nutrition, and physiotherapy, provide specialized expertise to enhance NCD care. By fostering collaboration among these disciplines, SRFCs could meet unmet needs in NCD care, providing holistic and innovative solutions for underserved communities.

Leveraging the SRFC model in Ghana presents a promising approach to addressing the country’s NCD crisis and, if successful, could serve as a model for the region. This study assesses stakeholder perspectives on the feasibility and support for implementing SRFCs in Ghana.

## Methods

## 1. Study Design and Setting

This study employed a mixed-methods exploratory sequential design to assess stakeholder interest in health professional student-led interventions for addressing access gaps in hypertension and diabetes management in Ghana (13). The research was conducted across two major public universities affiliated to teaching hospitals, Ghana Health Service (GHS) institutions, and nearby communities. The study unfolded in two phases: a qualitative phase involving focus group discussions (FGDs) and key informant interviews (KIIs), followed by a quantitative phase featuring a structured survey. Insights from the qualitative phase informed the development of the survey instrument to examined students’ motivation, perceived impact, and feasibility of the SRFC model.

Student and faculty interviews were conducted at the University of Ghana Medical School and the University of Cape Coast School of Medical Sciences. The University of Ghana, located in Accra, is the nation’s largest university (14). It is affiliated with Korle Bu Teaching Hospital which serves 400,000 patients annually, primarily from Accra’s urban, peri-urban, and slum populations, but also across Ghana and West Africa (15). The University of Cape Coast, in a less urban city town, is affiliated with Cape Coast Teaching Hospital, a 400-bed facility serving the city and surrounding rural communities (16).

Interviews with GHS community nurses and district health directors were conducted at the Mfantseman Ghana Health Service District Directorate in Saltpond. Interviews with higher-level officials and policymakers were conducted at the Ministry of Health and the Office of the National Presidential Advisor on Health, both located in Accra. Community members FGDs were conducted at Oguaa Market in Cape Coast and Korle Gonno Townsquare in Accra.

## 2. Data Collection Procedures

### 2.1 Qualitative Techniques

Between February 8th, 2024, and April 19^th^, 2024, we conducted FGDs and KIIs to explore stakeholders’ perspectives on non-communicable diseases (NCDs), access barriers, and interest in student-led care models.

- Sampling Strategy: Participants were purposively selected from medicine, pharmacy, nursing, optometry, public health, and allied health programs. Faculty members, university deans, GHS officials, and community members from areas surrounding the medical schools were also included. Eligibility criteria included age ≥18 years and ability to provide informed consent. Vulnerable populations, such as minors, pregnant individuals, and those with impaired capacity, were excluded.
- Data Collection Summary:
- `o` 6 FGDs with 48 health professional students
- `o` 6 KIIs with university faculty (deans, lecturers)
- `o` 4 KIIs with GHS officials, including public health nurses, district directors and national level policymakers.
- `o` 2 FGDs with 12 community members in Cape Coast and Accra
- Procedures: All discussions used a semi-structured guide covering NCD knowledge, perceived access gaps, and attitudes toward student-run interventions. Interviews and FGDs were conducted in English or local languages (Twi/Fante), audio-recorded with permission, and transcribed verbatim. Translations were reviewed by bilingual team members for accuracy.

### 2.2 Quantitative Techniques

The quantitative phase involved a structured survey administered from April 24^th^ to April 30^th^, 2024, to assess students’ attitudes toward participating in student-run clinics and perceptions of NCD care.

- Sampling and Administration: The survey was distributed to 316 health professional students across both universities using an online questionnaire. Students were recruited via academic listservs, in-class announcements, and student organization outreach. Participation was voluntary and anonymous, with electronic consent obtained beforehand.
- Instrument Design: The survey included items derived from qualitative themes, structured around Likert-scale and multiple-choice questions assessing:
- `o` Familiarity with common NCDs
- `o` Perceived burden of common NCDs.
- `o` Barriers to care and early screening.
- `o` Interest in leading or participating in SRFCs.

## 3. Data Analysis

### 3.1 Qualitative Phase

Thematic analysis was performed using NVivo (Release 1.7.2, QSR International, 2022) (17) to identify recurring patterns and themes.

- Familiarization: Transcripts were prepared and reviewed in full by four researchers (B.F., E.B., A.B., B.A.).
- Initial Coding: Two researchers (B.F. and B.A.) independently coded transcripts using an inductive, data-driven approach.
- Codebook Development: Codes were discussed, refined, and compiled into a shared codebook.
- Theme Generation: Codes were grouped into broader categories reflecting key research domains such as NCD burden, access gaps, and interest in student-led models.
- Validation: Discrepancies were resolved through consensus, and thematic outputs were reviewed with select participants for member-checking.

### 3.2 Quantitative Phase

Survey data were analyzed using SPSS version 28 (18)

- Descriptive Statistics: Used to summarize participant demographics, NCD familiarity, and levels of interest in SRFC involvement.
- Bivariate Analysis: Chi-square tests were conducted to assess differences in students’ willingness to participate across academic programs and year levels.
- Multivariable Analysis: Logistic regression examined whether familiarity with NCDs predicted willingness to lead or participate in SRFCs.
- Statistical Threshold: A p-value of <0.05 was considered statistically significant.

#### Ethical and Safety Considerations

Ethical approval was obtained from the GHS (GHS-ERC:040/11/23) and Yale University (2000036204) IRBs. Verbal and written consent was obtained from all participants. Before consent forms were signed, participants were provided with an information sheet that was read and explained to them. Participants were informed that participation was entirely voluntary and that they could withdraw at any time without penalty. They were assured that their decision not to participate or to withdraw would not affect their treatment or participation in any other aspect of the study. All data were de-identified and securely stored. No compensation was provided, but refreshments were offered. Data ownership and potential conflicts of interest were disclosed.

## Results

### Health Professional Participant Demographics

Health professional students’ demographic characteristics for the qualitative and quantitative phases are summarized in Table 1 and Table 2, respectively. A total of 48 students participated in the qualitative focus group discussions, while 316 students completed the quantitative survey. The mean age across both cohorts was approximately 22 years, with the qualitative sample ranging from 17 to 27 years (SD = 2.49) and the survey sample from 17 to 36 years (SD = 3.11). Gender distribution was relatively balanced, with females comprising 45.8% of qualitative participants and 52.8% of survey respondents. Participants represented a range of health professional programs—including nursing, pharmacy, allied health sciences, medicine, and optometry—with nursing and pharmacy students being the most represented in the survey phase (25.9% and 25.6%, respectively). Students were distributed across all academic levels, from first-year (Level 100) to final-year (Level 600), providing a broad spectrum of perspectives across the educational continuum.

**Table 1.**
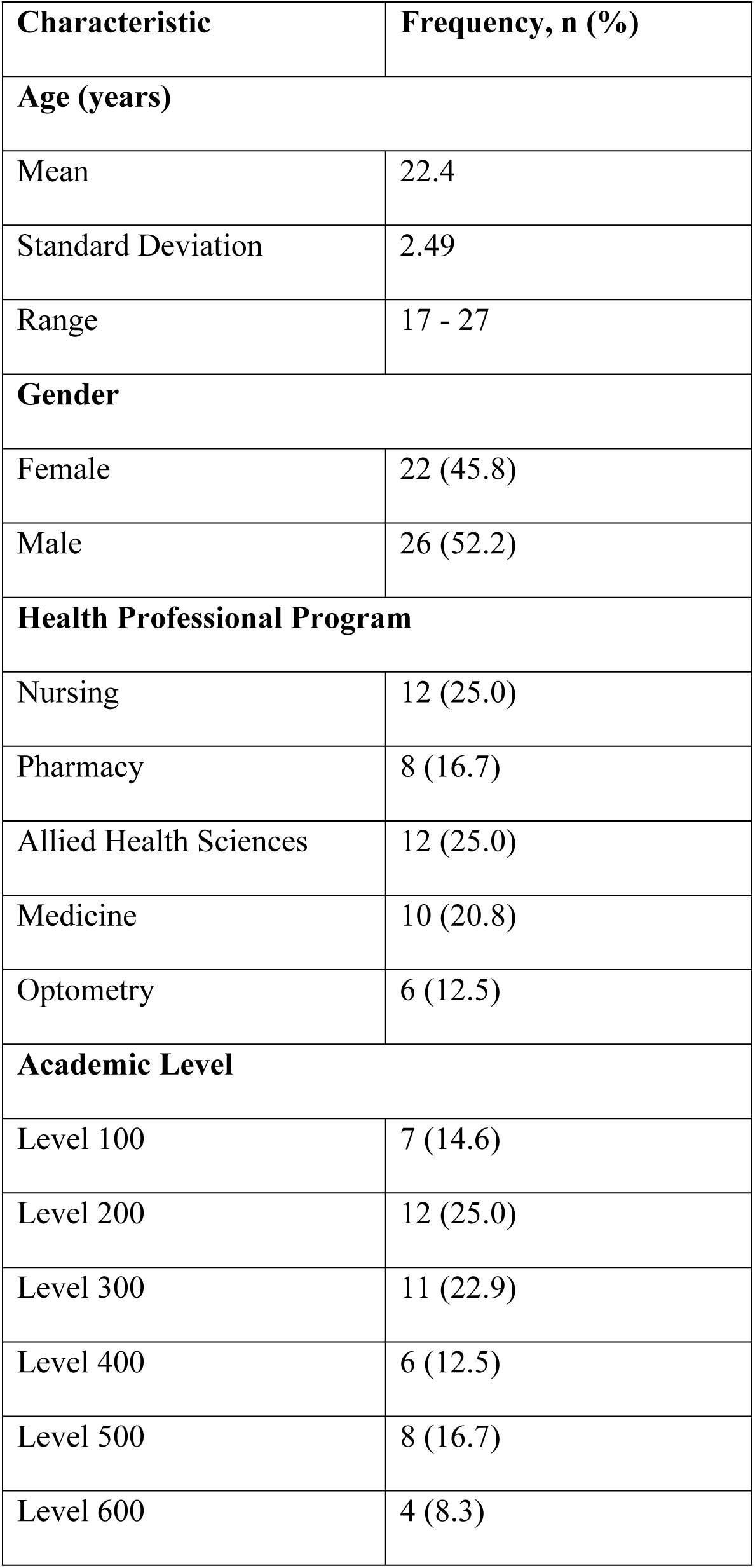
Descriptive statistics of demographics of FGD health professional student participants. **Demographic Characteristics of Health Professional Student FGD Participants (N = 48).**

**Table 2.** Descriptive statistics of health professional students’ demographics. **Demographic Characteristics of Health Professional Student Survey Respondents (N = 316).**

| **Characteristic** | **Frequency, n (%)** |
| --- | --- |
| **Age (years)** |  |
| Mean | 22.3 |
| Standard Deviation | 3.11 |
| Range | 17 - 36 |
| **Gender** |  |
| Female | 167 (52.8) |
| Male | 149 (47.2) |
| **Health Professional Program** |  |
| Nursing | 82 (25.9) |
| Pharmacy | 81 (25.6) |
| Allied Health Sciences | 72 (22.8) |
| Medicine | 55 (17.4) |
| Optometry | 17 (5.3) |
| Other | 9 (2.8) |
| **Academic Level** |  |
| Level 100 | 73 (23.1) |
| Level 200 | 65 (20.1) |
| Level 300 | 39 (12.3) |
| Level 400 | 64 (20.3) |
| Level 500 | 62 (19.6) |
| Level 600 | 13 (4.1) |

### Stakeholder Perspectives on SRFCs: Integrated Findings

Findings from both the qualitative and quantitative phases were integrated at the interpretation level using a narrative weaving approach. This mixed-methods integration allowed for a richer understanding of stakeholder perspectives on student-led responses to the growing burden of non-communicable diseases (NCDs) in Ghana. Across health professional students, university faculty, Ghana Health Service (GHS) officials, and community leaders, there was strong and consistent support for the concept of a Student-Run Free Clinic (SRFC) focused on NCD screening, early management, and self-care education.

Importantly, this support was shaped by both lived experiences and structural realities. Students not only demonstrated a deep familiarity with the landscape of NCDs in Ghana but also articulated the complex barriers that impede care—from sociocultural attitudes and low health literacy to systemic constraints like cost and infrastructure gaps. Their belief in the relevance of SRFCs was tied directly to how well the model aligned with these observed needs. At the same time, stakeholders across sectors recognized SRFCs as a feasible and complementary intervention—one that could extend the reach of existing health systems, support national priorities, and provide meaningful training opportunities for future health professionals.

To better understand the depth and contours of this support, three key domains emerged from our analysis: (1) students’ high level of familiarity with NCDs and perceived barriers to care; (2) strong acceptance of SRFCs as a viable and contextually appropriate intervention among students; and (3) widespread cross-sector endorsement from GHS, university leadership, and community stakeholders. These domains are explored in detail below.

### Key Domain 1: High Level of Familiarity with NCDs and Barriers to Care Among Students

Health professional students demonstrated a high level of familiarity with non-communicable diseases (NCDs), shaped by personal, academic, and clinical exposure. During focus group discussions, many students shared stories about family members, peers, or acquaintances affected by NCDs. They cited conditions such as hypertension, diabetes, cancer, and stroke, and described basic mechanisms of disease, long-term complications, and the broader health system impact. These reflections indicated both theoretical knowledge and lived experience.

*“My father had liver cancer but before that, he had diabetes, a terminal one. So, he passed away last year.” (University of Cape Coast Group 2 Student 2, UC2S2)*.

*“My roommate was living with diabetes. It’s not a good experience; trust me”. (UC1S5)*.

*“When I hear non-communicable diseases, what usually comes to mind is conditions like diabetes, hypertension, [] because they can’t be transmitted from one person to the other.” (University of Ghana Group 2 Student 3, UG2S3)*.

These qualitative findings were mirrored in the structured survey. A majority (58%) of respondents rated their familiarity with NCDs at level 4 or higher on a 5-point Likert scale (Figure 1), and the most frequently identified conditions matched those cited in FGDs (Figure 2). Furthermore, 93.4% (n = 295) of student respondents agreed that NCDs represent a major health issue in Ghana, highlighting strong awareness of the national disease burden.

**Figure 1.**
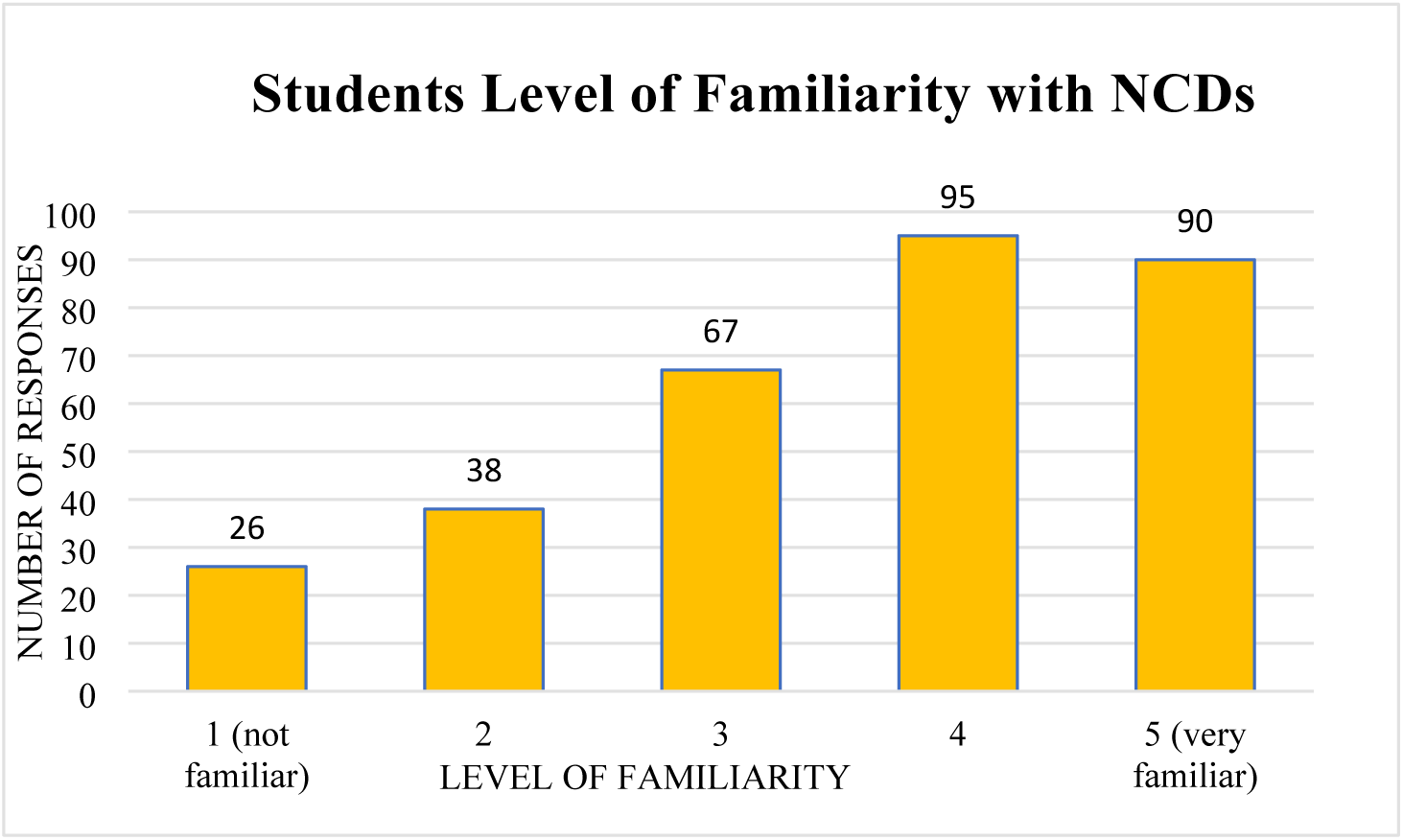
Health professional students’ rating of their familiarity with NCDs (1 = No familiarity; 5 = High familiarity).

**Figure 2.**
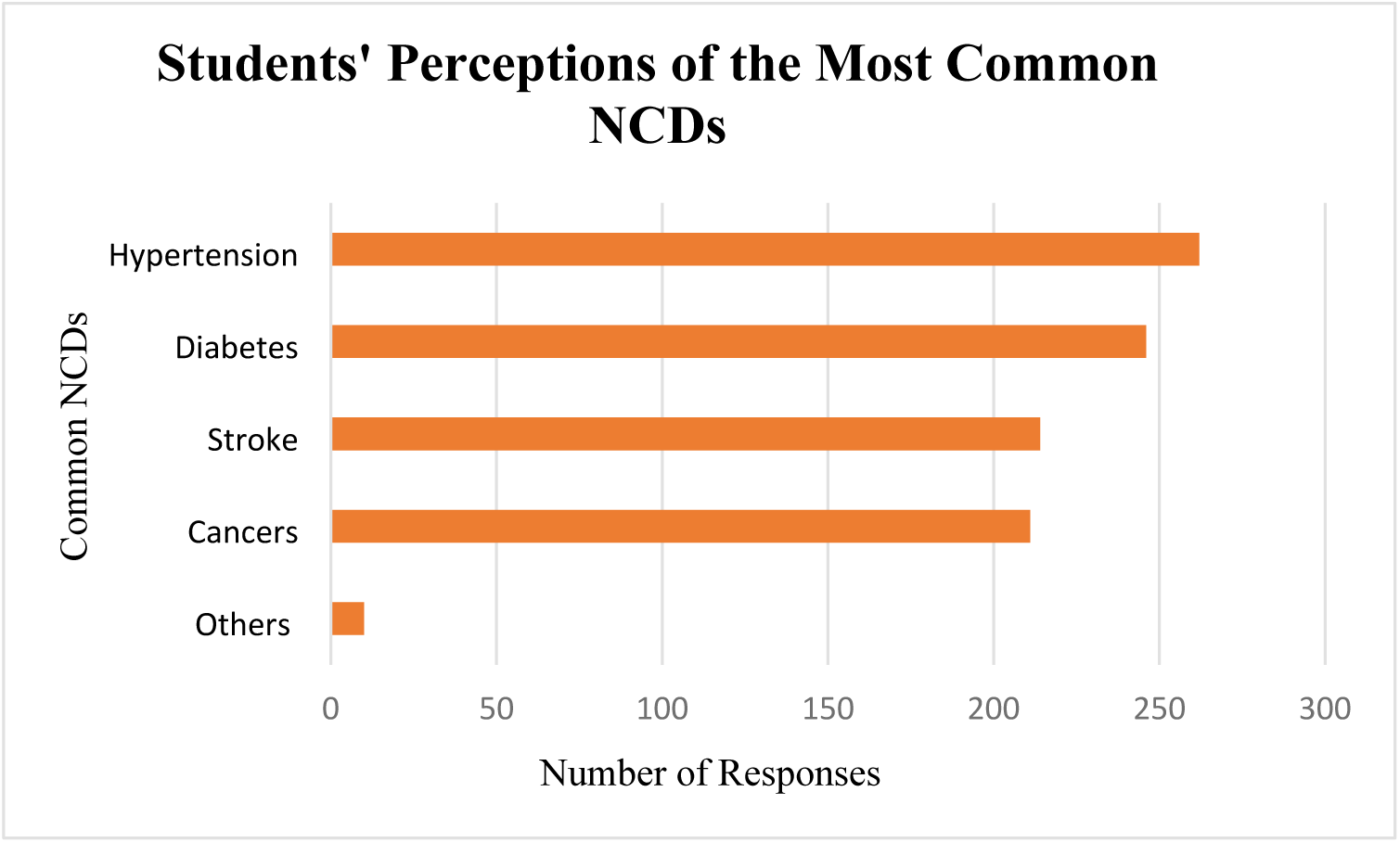
Health professional students’ perception of the most common NCDs. “Others” include asthma, sickle cell anemia, jaundice, and thyrotoxicosis.

In addition to disease familiarity, students demonstrated a nuanced understanding of barriers to NCD care. Thematic analysis of FGDs revealed four core areas shaping these perceptions: limited awareness and understanding, socio-cultural influences, health system limitations, and financial constraints. Students frequently described community-level misconceptions about NCDs and emphasized how the asymptomatic nature of many conditions reduced urgency to seek care. Religious interpretations and cultural attitudes were also said to discourage early intervention. Structural challenges—including inadequate facilities, understaffing, and weak follow-up mechanisms—were consistently cited. Finally, the cost of diagnostic testing and chronic care emerged as a critical barrier, particularly for low-income households.

Table 3 summarizes these five themes and presents illustrative student quotes from FGDs.

**Table 3.** Summary of Student Perspectives on Barriers to NCD Care. **Summary of Student Perspectives on Barriers to NCD Care.**

| Themes from FDGS | Illustrative Student Quotes |
| --- | --- |
| Limited Awareness and Understanding of NCDs | *“Most of the market [women], when their head is aching, they just walk in [to a drug store] and they get [paracetamol]... that headache could be anything.” (UG2S2)*<br>*“Sometimes they know, but they don’t know much... they just know the surface like ‘I have high blood pressure.’” (UG1S3)* |
| Socio-Cultural Barriers | *“Some people are strongly attached to their religious affiliations... their pastors tell them, ‘As a child of God, the Holy Spirit will heal you.’” (UG2S6)*<br>*“They are indifferent about it... they say, ‘Is it because we will die that we won’t sleep?’” (UC3S3)* |
| Health System Limitations | *“Even if they have a CHPS compound, they don’t have health professionals there... just one nurse or something.” (UC3S2)*<br>*“The follow-up system in Ghana is not proper... sometimes they call once or twice and give up.” (UG1S3)* |
| Financial Constraints | *“Most people would not prefer to visit the hospitals because of the rate at which bills are being increased.” (UC2S3)*<br>*“Let’s say the drug costs 50 cedis, but the person earns only 5 cedis a day... and came with just 10 cedis. How do you expect them to buy it?” (UC1S2)* |

**Table 3.** Association between academic program and willingness to participate in an SRFC (χ² = 10.25, p = 0.069). **Academic Program Vs. Willingness to Participate.**

| Academic Program | Yes, n (%) | No, n (%) |
| --- | --- | --- |
| Allied Health Sciences | 60 (83.3) | 12 (16.7) |
| Nursing | 65 (79.3) | 17 (20.7) |
| Pharmacy | 75 (92.6) | 6 (7.4) |
| Medicine | 51 (92.7) | 4 (7.3) |
| Optometry | 16 (94.1) | 1 (5.9) |
| Other | 7 (77.8) | 2 (22.2) |
| Total | 274 (86.7) | 42 (13.3) |
| Chi-Square ($\chi^2$) | 10.25 | |
| P-value | 0.069 |  |

**Table 4.** Association between academic level and willingness to participate in an SRFC (χ² = 3.77, p = 0.583). **Academic Level Vs. Willingness to Participate.**

| Academic Level | Yes, n (%) | No, n (%) |
| --- | --- | --- |
| Level 100 | 63 (86.3) | 10 (13.7) |
| Level 200 | 57 (87.7) | 8 (12.3) |
| Level 300 | 34 (87.2) | 5 (12.8) |
| Level 400 | 56 (87.5) | 8 (12.5) |
| Level 500 | 55 (88.7) | 7 (11.3) |
| Level 600 | 9 (69.2) | 4 (30.8) |
| Total | 274 (86.7) | 42 (13.3) |
| Chi-Square ($\chi^2$) | 3.77 | |
| P-value | 0.583 |  |

**Table 5.** Logistic regression: familiarity with NCDs as a predictor of SRFC participation (OR = 1.41, p = 0.007). **Familiarity with NCDs vs. Willingness to Participate.**

| Parameter | Coefficient | P-value | Odds Ratio |
| --- | --- | --- | --- |
| Intercept | 0.7058 | 0.1102 | 2.0254 |
| Familiarity with NCDs | 0.3454 | 0.0071 | 1.4126 |

### Key Domain 2: Students Acceptance of SRFCs as an Appropriate NCD Intervention

Health professional students expressed strong interest in participating in a Student-Run Free Clinic (SRFC) focused on non-communicable disease (NCD) screening, early management, and self-care education. This interest was evident in focus group discussions and reinforced by survey findings, where 274 out of 316 respondents (86.7%) indicated willingness to participate in such an initiative.

Chi-square tests revealed no statistically significant association between willingness to participate and students’ academic program (p = 0.069) or academic level (p = 0.583), indicating broad support across different disciplines and training stages. However, logistic regression analysis identified familiarity with NCDs as a significant predictor of willingness to participate (β = 0.345, p = 0.007), with each unit increase in NCD familiarity associated with a 41% increase in the odds of participation (odds ratio = 1.41). These results suggest that students’ interest in SRFCs is widespread and not confined to a specific training group—but that greater familiarity with NCDs further enhances their likelihood of engagement.

Students’ strong willingness to participate was further supported by their perceptions of SRFCs as an effective intervention for addressing key barriers to NCD care. When asked to rate the effectiveness of SRFCs across screening, early management, and education on a 5-point scale, most respondents selected 4 or 5. Specifically, 197 students (62%) rated SRFCs as highly effective in improving NCD screening (Figure 3), and 225 students (71%) provided similar ratings for educational effectiveness, with 109 giving the maximum score (Figure 5). Perceptions of early management effectiveness also remained consistently high (Figure 4).

**Figure 3.**
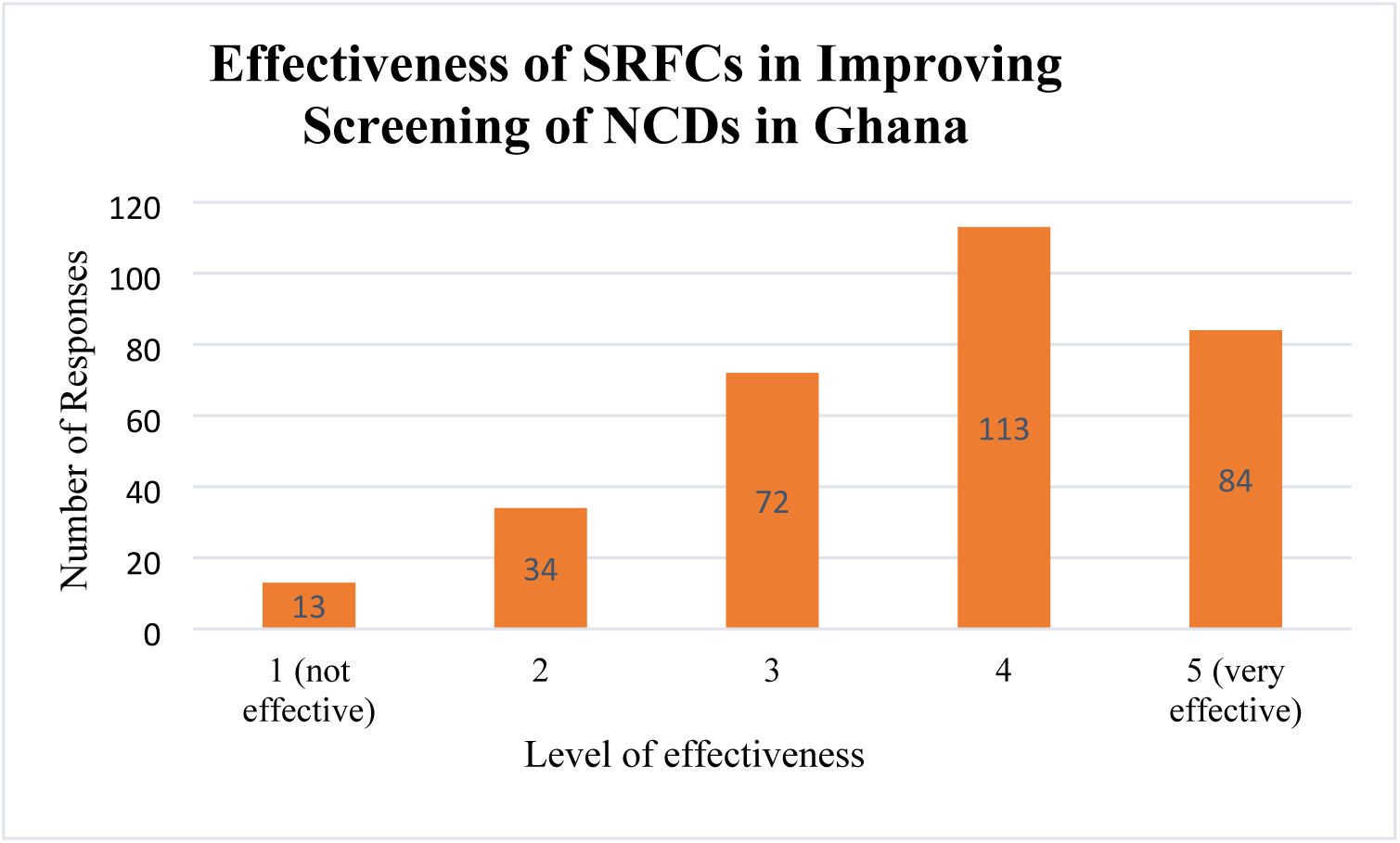
Health professional students’ perception of the effectiveness of SRFCs in improving screening of NCDs in Ghana. Level 1 = Not Effective; Level 5= Very Effective.

**Figure 4.**
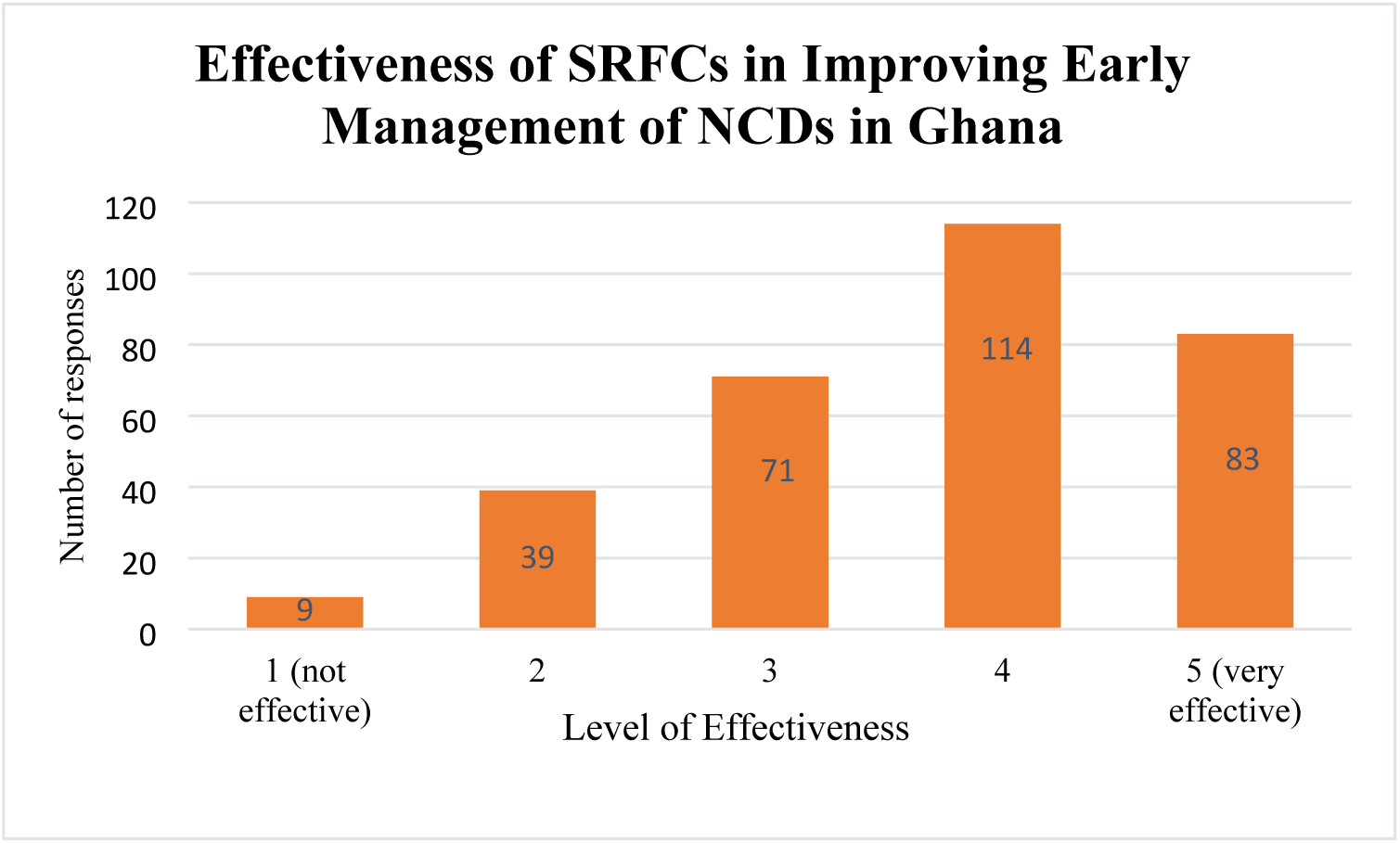
Health professional students’ perception of the effectiveness of SRFCs in improving early management of NCDs in Ghana. Level 1 = Not Effective; Level 5= Very Effective.

**Figure 5.**
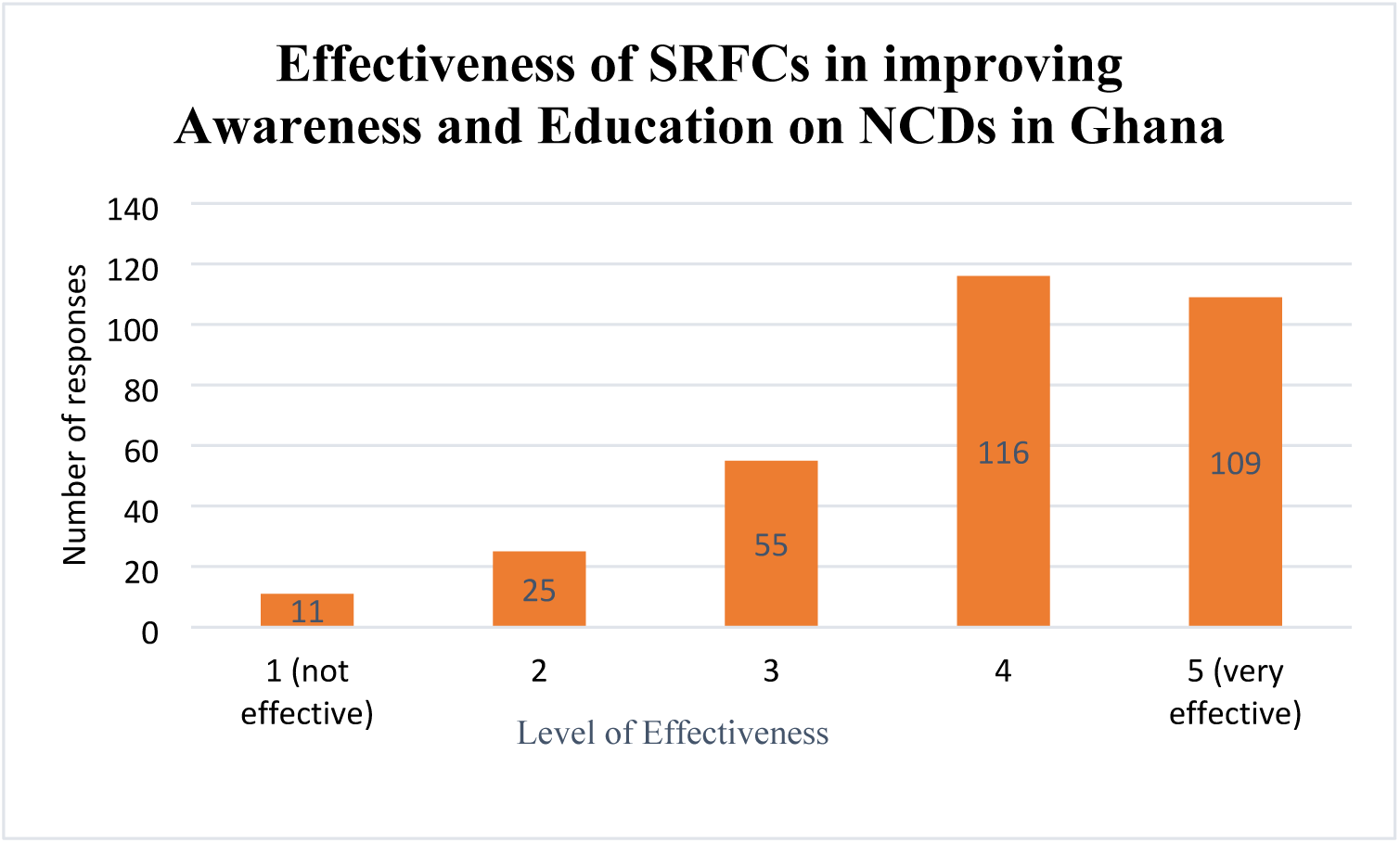
Health professional students’ perception of the effectiveness of SRFCs in improving education of NCDs in Ghana. Level 1 = Not Effective; Level 5= Very Effective.

These perceptions were further supported by students’ views on the specific barriers SRFCs are best positioned to address. As shown in Figure 6, students most frequently identified the high cost of healthcare services, lack of healthcare facilities, and limited education and awareness of NCDs as the top barriers SRFCs could effectively mitigate. Notably, these align directly with the top three barriers elicited earlier in the study (see Figure 1), suggesting a strong convergence between perceived need and perceived effectiveness. This alignment indicates that students viewed SRFCs not only as a broadly useful model, but as a reliable and context-specific intervention for addressing the most critical gaps in NCD care in Ghana.

**Figure 6.**
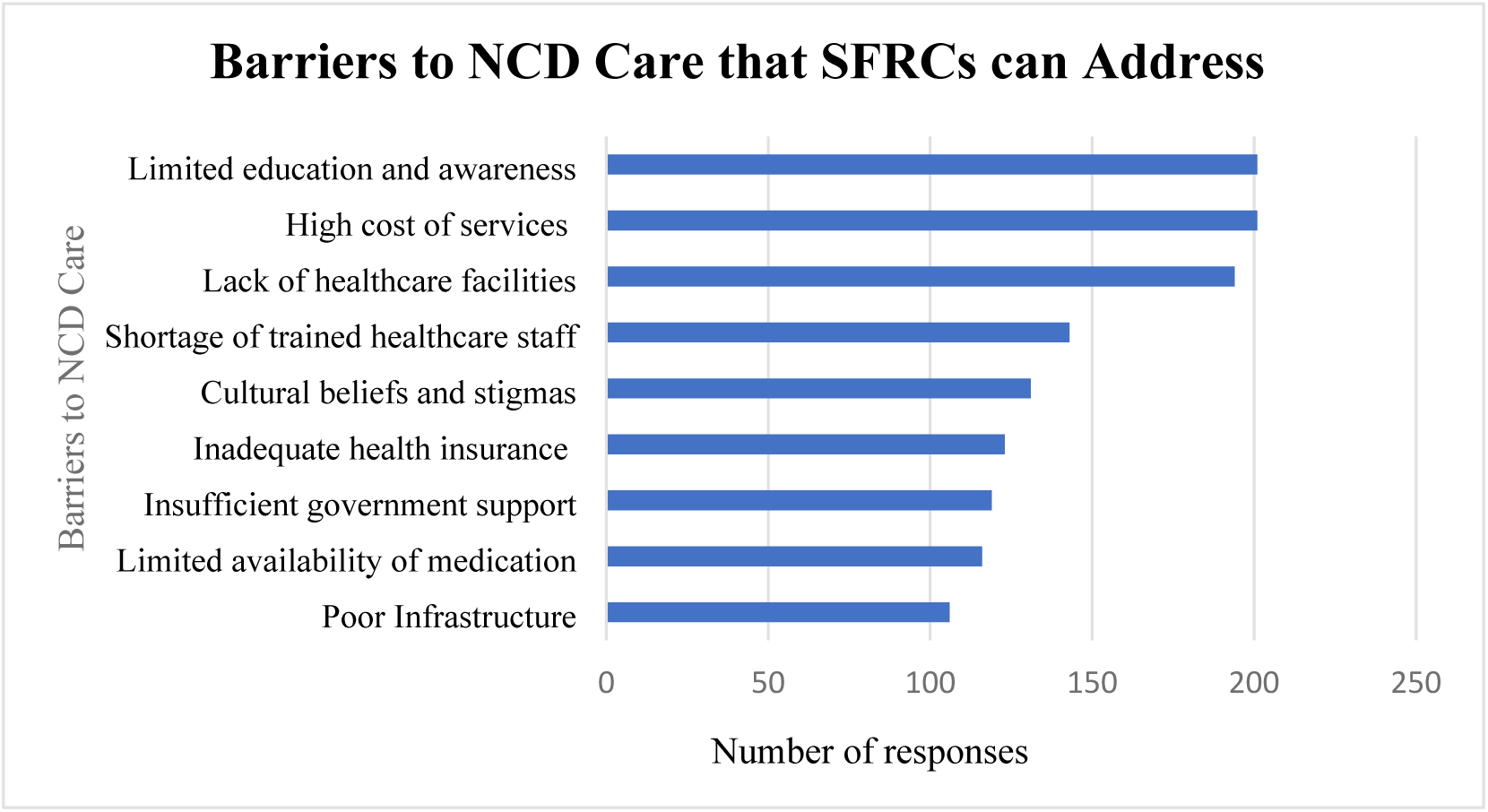
Barriers to NCD screening, early management, and self-care education that SRFCs can address.

Focus group discussions provided deeper insights into the motivations driving student interest in SRFCs, as well as the challenges that might hinder participation. Students frequently cited their desire to serve communities and improve access to care for underserved populations. They also valued SRFCs as opportunities for hands-on learning, clinical skill development, and interprofessional collaboration. Some noted that logistical or symbolic incentives—such as mentorship, recognition, or refreshments—could further encourage participation, especially when balanced with academic demands.

However, students also identified notable barriers. The most frequently cited was academic workload, with concerns that participation could conflict with lectures, exams, or rotations. Students also raised concerns about the personal cost of transportation to clinic sites and the availability of essential materials. Finally, safety concerns—particularly about traveling to unfamiliar or insecure areas—were mentioned as a potential deterrent.

| Category | Theme | Illustrative Quotes |
| --- | --- | --- |
| Motivating Factors | Community and Social Impact | *Yeah, I absolutely would. It's great to give back to the community. And it looks good on the resume. It gives me experience in the clinic as well.” (UG2S5)* |
|  | Academic and Professional Development | *“Personally, I would also join if there is an opportunity because one, it will help me to learn and two, help me to build connections, not only with the health professionals but also with the students and [] the patients that will come.” (UC2S4).*<br>*“I think it's a great opportunity that should be implemented.[] it gives us another opportunity to further our skills.” (UC2S1).* |
|  | Opportunities for Interprofessional Collaboration | *“I have not gotten the chance to I mean attend program with different health professionals. So even seeing that people from different health areas are coming together for such a program like this, I think it will be wise to be a part of this so that we all share knowledge and learn from each other” (UC1S7).* |
|  | Incentives | *“Students want ‘item 13’ [refreshments] and T-shirts. So, if you provide that, I will keep showing up”. (UG3S2).* |
| Discouraging Factors | Academic Obligations and Conflicts | *“Okay, for me personally, if it doesn’t clash with any of my academic activities, why not? Because I am enthused about helping.” (UC2S1)*<br>*“I feel I’ll be interested but maybe like I was saying, if it’s going to against my academics or I’m going to get stressed out or something, I’m sure I’ll drop out at some point”. (UG1S3)* |
|  | Resource Demands | *If there is no means of transport and I will have to spend to go there, I won't be that happy to go for the thing. I will be reluctant. As well as availability of resources to work with” (UC2S7)* |
|  | Safety Concerns | *“I will also say the security. [] I wouldn't go to a place where I know they can attack me anytime. Certain areas when you go there you know they can attack you” (UC2S5).* |

### Key Domain 3: Cross-Sector Endorsement of SRFCs as a Contextually Appropriate Response to NCD Care Gaps

Stakeholders across the Ghana Health Service (GHS), university leadership, and community groups consistently endorsed Student-Run Free Clinics (SRFCs) as a relevant and timely intervention for addressing key gaps in non-communicable disease (NCD) care in Ghana.

Despite representing distinct institutional and social roles, participants expressed overlapping motivations rooted in a shared commitment to expanding healthcare access, investing in student capacity, and strengthening partnerships between health systems, universities, and communities. This cross-sector endorsement was reflected across three convergent subdomains: alignment with institutional and community goals, confidence in students’ ability to deliver services, and recognition of SRFCs as valuable platforms for student training and interprofessional collaboration.

### A. Alignment with Institutional and Community Goals to Expand Access

Stakeholders from GHS and universities emphasized that SRFCs align with their broader mandates to serve underserved communities and promote health equity. Faculty members framed SRFCs as natural extensions of existing outreach programs—such as the Community-Based Experience and Service (CoBES)—where students already engage in public education, disease monitoring, and referrals under supervision.

*“UCC is known for its community service. For instance, at the School of Pharmacy, we do CoBES, where students go into the community and then, yes, render healthcare services. Some of the things they do is to educate the public, and again, to monitor some of these diseases, and do the referrals where appropriate. [] So, on our standards, we should encourage [any student-led initiative]” (UCF3)*.

GHS officials similarly viewed SRFCs as compatible with national health promotion efforts and existing community infrastructure, particularly the Wellness Clinics. They highlighted a foundation of goodwill between GHS and local communities that student-led efforts could readily tap into, noting that official endorsement would enhance credibility and facilitate community buy-in.

*“When we mention our health promotion efforts, it’s necessary to mention that we love to collaborate. So, by all means, we’ll collaborate because one cannot do the work all alone. Students can partner with the Wellness Clinics and support the work we do. That will surely increase access for so many people.” (Ghana Health Service Official 2, GHS2)*

*“The Ghana Health Service freely goes out to areas like the rural settings and in fact the district zones…So already, the goodwill is there, all you need is to just tap into it.” (GHS4)*

*“The goodwill support is guaranteed. On our end, we can let the communities know that this clinic has our blessing and their universities’. Any push we can give, we will.” (GHS2)*

Community members echoed this urgency and enthusiasm, describing SRFCs as critical responses to unmet needs in screening, education, and early management of NCDs.

*“These diseases are killing us, but we don’t know. So, if “little doctor” can tell us, do this, don’t do that and they can explain the reason to us. It will help” (Community Reception Interview 2 Participant 3, CR2P3)*.

*“When this initiative is established and we go for screening and some discoveries are uncovered, we will then be able to address it. But until then, we won’t know. We need this very urgently.” (CR1P1)*

### B. Confidence in Student Capacity and Community Trust

Faculty members and GHS officials described students as capable contributors to public health outreach. They cited prior examples of student-led initiatives, like the annual “Health Weeks” where students lead several social and communal health initiatives, as evidence of their leadership and organizational potential. They emphasized that under appropriate supervision, students could extend NCD services and promote early detection through community engagement.

*“We believe students can contribute to identifying these disease conditions early and also promote health awareness through health education, outreaches to let the community-dwelling citizens be aware that these are conditions that are mainly manageable and could prevent complications.” (UGF1.)*

*“We have a week set aside, we call it the health week, where students on their own are sent to different regions and different areas of the country. Usually, we come up with a theme, whether it’s Malaria prevention or hypertension prevention and then the students go out and contribute through community engagement activities in line with the theme of the year. (UGF2)*.

*Students can help in the normal health screening like the BP check, [for] diabetes, the blood sugar check, they can do the fasting blood sugar checks and all those things”. (GHS3)*

GHS personnel reinforced this confidence, noting that they already supervise trainees and would be willing to support student efforts in SRFCs through on-site mentorship and clinical guidance.

“We know these students already and we know what they can do. So, when they come, we can guide them with what they cannot do. With the simple tasks like blood glucose screening, BP checks, and the education, we can supervise them. We do that already.” *(GHS1)*

Community members also cited positive interactions with students in previous initiatives as a reason for their support. Many had built trust with students through past experiences and looked forward to future engagements. Some shared anecdotes of assisting students with tasks like setting up clinic canopies and spreading awareness through both official announcements and word of mouth. They appreciated students’ patience, attentiveness, and willingness to listen—qualities that made care feel truly patient-centered and fostered a strong sense of connection.

*“Whenever these students come around, we help them assemble the canopies. They have time and take care of us, and this has been very helpful for us. Through this, we’ve been able to establish a sort of connection with these students as well.” (CR1P6)*

### C. Value for Student Training and Interprofessional Collaboration

University faculty highlighted that student-led initiatives provide essential hands-on training, enhancing clinical knowledge, patient interaction skills, and workforce readiness. They emphasized the alignment of student-led initiatives with interprofessional education goals, citing ongoing efforts to integrate interprofessional courses into the curriculum. They noted that students also recognized the benefits of collaboration, especially in community outreach.

*“It is a very good idea. It is something everybody should embrace and it’s something we should all encourage. Last year, if I may remember, the College [of Health and Allied*

*Sciences] started trying to put up interprofessional courses and programs. [] It’s something we are looking at and we know the advantages it will offer the students”. (UCF3)*.

*“Yes, I was in a particular meeting for instance, and that meeting, I think the medical student brought up those ideas [for interprofessional collaboration]. They [went] to the community, and they realized that if the nurses were there, it would have been a great experience”. (UCF2)*.

Faculty also highlighted SRFCs as opportunities for junior students to learn from more advanced peers in real-world settings.

*“We also believe that it’s an opportunity for some of our students in clinical years to also be there to understudy their senior colleagues.” (UCF2)*.

Community members also viewed the clinic as a crucial part of students’ education and were eager to support it. They recognized the importance of hands-on training in preparing students for future healthcare roles, understanding that these students would one day be responsible for their care.

*“These students are coming to practicalize what they have been taught in taking care of us. And if these students are not given the requisite practical training, how then will they be able to effectively go about with their future career which entails taking care of us? So, for me, I am not bothered at all.” (CR1P3)*

## Discussion

This study highlights broad stakeholder interest in student-led clinical interventions, such as SRFC, for NCD care in Ghana, emphasizing their potential to improve healthcare access in underserved communities. The findings align with existing literature on the role of SRFCs in expanding primary care services while simultaneously enhancing student training and interprofessional collaboration. The strong familiarity with NCDs and their associated barriers among stakeholders underscores the relevance of such interventions, particularly in addressing gaps in early detection, management, and community education.

Institutional alignment further supports the feasibility of SRFCs, with university faculty and GHS officials recognizing their potential to complement existing health services. Notably, faculty emphasized that students have successfully participated in previous community health initiatives, reinforcing confidence in their ability to contribute meaningfully to NCD care. The strong student enthusiasm, with 86.7% expressing willingness to participate, suggests that such initiatives could be sustained through ongoing student engagement. The identified opportunities for interprofessional collaboration are particularly promising, as interdisciplinary teamwork has been shown to improve health outcomes and enhance educational experiences.

This research contributes to the growing body of literature on SRFCs, particularly in low-resource settings like Ghana, where limited studies have explored their feasibility. While SRFCs are well-documented in high-income countries, particularly in the United States, their implementation in Sub-Saharan Africa remains underexplored (11,19). Although SHAWCO at the University of Cape Town serves as a longstanding model, its establishment occurred under vastly different health and regulatory conditions (2). Additionally, existing SRFC frameworks in high-income countries differ significantly from those in low-resource settings like Ghana. In high-income contexts, SRFCs are typically embedded within well-resourced academic institutions, benefit from clear supervisory structures, legal protections, and access to diagnostic tools. In contrast, Ghana faces limited clinical infrastructure, faculty shortages, and unclear policies on student-led care. These differences highlight the need for localized feasibility assessments to ensure safe and context-appropriate implementation.

By highlighting the role of interprofessional collaboration among students from various health disciplines, this study also builds on existing literature that emphasizes the importance of team-based care in managing complex conditions (20,21). However, the literature often focuses on professionalized teams, and this research uniquely explores the feasibility of such collaboration at the student level. Despite the numerous advantages and push for interprofessional education (IPE) and training by organizations such as the Africa Interprofessional Education Network, IPE is still not well established in low- and middle-income countries, particularly across Sub-Saharan Africa, where the burden of disease is greater (22,23). Thus, this study provides additional evidence for the interest and need for an interprofessional collaboration among health professional students. Additionally, the study’s focus on the community-based setting of SRFCs addresses a critical gap in understanding how such initiatives can be tailored to local contexts, where access to formal healthcare services is often limited.

### Implications for Policy and Practice

These findings provide valuable insights for policymakers, educators, and healthcare leaders considering the integration of student-led clinics into Ghana’s healthcare landscape. First, formalizing partnerships between universities, the GHS, and community health facilities could provide a structured framework for SRFCs, ensuring alignment with national health priorities. Second, incorporating supervised SRFC activities into medical and health professional training curricula could enhance hands-on learning while addressing service delivery gaps. Finally, clear guidelines for interprofessional collaboration could further strengthen SRFCs, enabling students from different health disciplines to work together effectively in community settings.

### Strengths and Weaknesses

This study offers several strengths, including its mixed-methods approach, which combines qualitative insights from key stakeholders with quantitative analysis of student perspectives. The inclusion of diverse voices—from faculty to community members—enhances the depth and applicability of the findings. Additionally, the study contributes novel data on the feasibility of SRFCs in Ghana, where limited research exists on this model.

However, some limitations should be acknowledged. The study relied on self-reported data, which may introduce social desirability bias, particularly regarding students’ willingness to participate. Additionally, while the sample included key institutional and community stakeholders, broader engagement with policymakers and healthcare administrators could further strengthen the findings. Future research should explore long-term implementation strategies, sustainability mechanisms, and the direct impact of SRFCs on health outcomes in Ghana.

### Conclusion

SRFCs present a promising model to address NCD care gaps in Ghana and, in turn, across Africa, while simultaneously providing valuable training opportunities for health professional students. Strong institutional support, demonstrated student capacity, and community buy-in suggest that a high degree of feasibility of SRFC in this context. Future research should remain stakeholder-engaged and should explore contextual adaptation of the SRFC model with a focus on implementation strategies and potential funding options to support the model locally.

## Data Availability

All relevant data are within the manuscript. De-identified survey datasets and qualitative excerpts supporting the findings are available upon reasonable request from the corresponding author.

## Competing Interests

The authors have declared that no competing interests exist.

## Funding Statement

This research received funding from the Richard A Moggio Fund, Office of Student Research, Yale School of Medicine.

## Author Contributions

**Brian A. Fleischer**: Conceptualization (equal); Data curation (lead); Funding acquisition (lead); Formal analysis (lead); Investigation (equal); Methodology (equal); Project administration (lead); Writing – original draft (lead); Writing – review & editing (equal)

**Jeremy I. Schwartz**: Conceptualization (equal); Funding acquisition (supporting); Methodology (equal); Supervision (equal); Validation (equal); Writing – review & editing (equal)

**Esi Berkoh**: Data curation (supporting); Investigation (equal); Writing – review & editing (equal)

**Bismark Amoh**: Formal analysis (supporting); Investigation (equal); Writing – review & editing (equal)

**Afriyie Badu**: Investigation (equal); Writing – review & editing (equal)

**Derek A. Tuoyire**: Methodology (supporting); Project administration (supporting); Supervision (equal); Validation (equal); Writing – review & editing (equal)

## Appendix List of Abbreviations

CHOs: Community Health Workers
CHPs: Community-based Health Planning and Services
CVDs: Cardiovascular Diseases
NCD: Non-Communicable Diseases
NHIS: National Health Insurance Scheme
NOP: Networks of Practice
NPHWs: Non-Physician Healthcare Providers (NPHWs)
SHAWCO: Students’ Health and Welfare Centers Organization
SRFC: Student Run Free Clinic

## Notes

### Competing Interest Statement

The authors have declared no competing interest.

### Funding Statement

This work was supported by the Richard A. Moggio Fund through the Office of Student Research at Yale School of Medicine. The funder had no role in study design, data collection and analysis, decision to publish, or preparation of the manuscript.

### Author Declarations

Ethical approval was obtained from the GHS (GHS-ERC:040/11/23) and Yale University (2000036204) IRBs

